# Using linear and natural cubic splines, SITAR, and latent trajectory models to characterise nonlinear longitudinal growth trajectories in cohort studies

**DOI:** 10.1101/2021.05.26.21257519

**Authors:** Ahmed Elhakeem, Rachael A. Hughes, Kate M. Tilling, Diana L. Cousminer, Stefan A. Jackowski, Tim J. Cole, Alex S.F. Kwong, Zheyuan Li, Struan F.A. Grant, Adam D.G. Baxter-Jones, Babette S. Zemel, Deborah A. Lawlor

## Abstract

Longitudinal data analysis can improve our understanding of the influences on health trajectories across the life-course. There are a variety of statistical models which can be used, and their fitting and interpretation can be complex, particularly where there is a nonlinear trajectory. This paper provides a guide to describing nonlinear growth trajectories for repeatedly measured continuous outcomes using linear mixed-effects (LME) models with linear splines and natural cubic splines, nonlinear mixed effects Super Imposition by Translation and Rotation (SITAR) models, and latent trajectory models. The underlying model for each of the four approaches, the similarities and differences between models, and their advantages and disadvantages are described. Their applications and correct interpretation are illustrated by analysing repeated bone mass measures across three cohort studies with 8,500 individuals and 37,000 measurements covering ages 5-40 years. Linear and natural cubic spline LME models and SITAR provided similar descriptions of the mean bone growth trajectory and growth velocity, and the sex differences in growth patterns. Latent trajectory models identified up to four subgroups of individuals with distinct trajectories during adolescence and similar trajectories in childhood and adulthood. Recommendations for choosing a modelling approach are provided along with a discussion and signposting on further modelling extensions for analysing trajectory exposures and outcomes, and multiple cohorts. In summary, we present a resource for characterising nonlinear longitudinal growth trajectories, that could be adapted for other complex traits. Scripts and synthetic datasets are provided so readers can replicate trajectory modelling and visualisation using the open-source R software.

## INTRODUCTION

Appropriate modelling of repeated measures in cohort studies can improve our understanding of: patterns of change across the life-course (e.g., developmental trajectories to peak function and age-related decline); (ii) influences on these patterns of change; and (iii) influence of variation in patterns of change on later health and wellbeing [1]. Many developmental processes display non-linear patterns of change with age, especially during the growing years, which makes it important but challenging to accurately model their trajectories [2]. Also requiring attention is the choice of method to appropriately address the research question, e.g., whether to use methods that model an average trajectory in the whole sample [3-5] or clustering based approaches to identify groups of individuals with similar trajectories [6]. An overview of sophisticated modelling procedures with open-source software applications in different cohorts is needed to address these challenges.

This paper provides a guide to describing nonlinear longitudinal growth trajectories for a single repeatedly-measured continuous outcome using linear and natural cubic splines [3, 4], SITAR (Super Imposition by Translation and Rotation) models [5], and latent trajectory models [6] – all common methods for examining growth. The following section gives an overview of modelling nonlinear growth and the various models considered. The four approaches (and appropriate interpretation of results) are illustrated by modelling bone mass trajectories across three cohort studies to characterise patterns of change and differences between male and females. The final section provides recommendations about when different modelling methods might be useful and discusses extensions and challenges in analysing exposures and outcomes of patterns of change and making cross-cohort comparisons.

## MODELLING NONLINEAR GROWTH

A variety of statistical methods are available for handling repeated (correlated) observations from the same individuals and analysing trajectories [7]. Most methods involve fitting growth models within either a structural equation modelling framework (e.g., latent growth curve analysis where change is analysed as a latent process [8]) or a multilevel modelling framework, with both giving similar results under certain conditions [9]. One type of multilevel model that can be useful if the primary interest is estimating a population-average trajectory is generalised estimating equations (GEE) [10, 11]. This uses a working covariance matrix to correct for dependence among repeated observations, and usually not suited for examining variation within/between individuals. Another multilevel model that can estimate both population-average and individual-specific trajectories (and which is more robust to missing outcome data than GEE) is the mixed-effects model [12].

### Mixed-effects models

Mixed-effects models (random-effects, multilevel, or hierarchical models) estimate a population-average trajectory as ‘fixed effects’ and variation of individual trajectories around this average as ‘random effects’ [11-13]. A common form is the linear mixed-effects (LME) model where the repeated outcome is modelled by a linear combination of the fixed and random effects. For a single continuous outcome (e.g., weight), an LME model for the outcome, as a linear function of time, which includes random intercepts and random slopes can be written as follows:

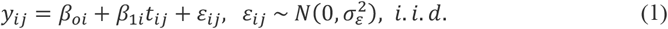

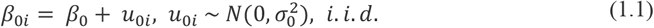

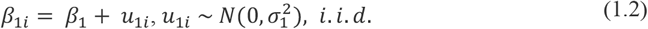

where, *y*_*ij*_ denotes a single outcome *y* measured in individual *i*(*i*= 1, 2, …, *N*) at time *t*_*ij*_ (*j* = 1, 2, …, *J*_*i*_), with responses (*y*_1_ … *y*_*N*_) assumed to be independent between individuals*β*_*oi*_ and *β*_1*i*_ are individual-specific intercept and slope terms (respectively) that have fixed effects (*β*_*o*_, *β*_1_) and random effects (*u*_*oi*_, *u*_1*i*_). The random effects *u*_*i*_ are assumed to be independently normally distributed with mean zero and covariance matrix *Ω*_*u*_. Residual errors *ε*_*ij*_ are assumed to be independently identically normally distributed with variance 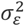 and reflect the difference between observed and predicted values for individual *i*at occasion *j*. The random effects *u*_*i*_ and residuals *ε*_*ij*_ are assumed to be mutually independent.

### Moving beyond a linear trajectory

The LME model in (1) assumes linear change in the outcome with time (e.g., age). Nonlinear change, which is common (particularly when modelling change over a large age range) can be incorporated into LME models by including linear combinations of nonlinear terms for age in the model – i.e., keeping the linear link function. Historically, the standard approach has been to use polynomial functions to approximate nonlinear curves. However, as in **Fig. 1**, polynomials have limitations, e.g., simpler polynomials give few curve shapes and more complex polynomials tend to fit badly at extremes and produce artefactual turns in the curve. A more flexible alternative to modelling complex patterns of change (e.g., with several peaks and troughs, as with body mass index (BMI) over infancy, childhood, and adolescence) is using spline functions.

**Fig. 1.**
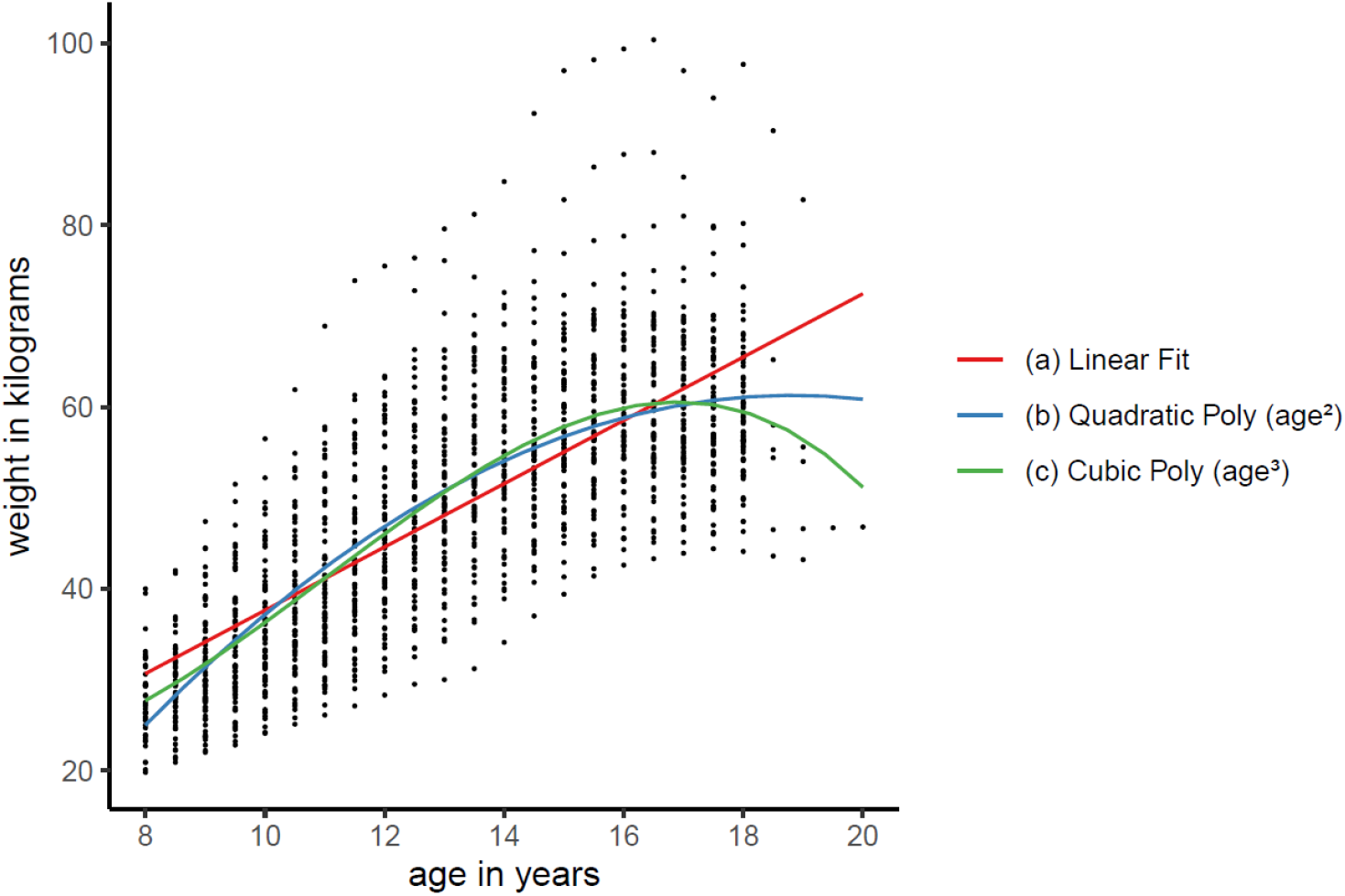
Example illustrating the limitations of using polynomial functions to approximate a nonlinear growth trajectory. Coloured lines represent predicted trajectories from LME models with age as (a) linear term and as (b) quadratic polynomial and (c) cubic polynomial. Points display weight measurements taken from 70 females in the Berkeley Child Guidance Study. Dataset was originally provided as an appendix to the book by Tuddenham and Snyder (1954). The data used in this example were taken from the freely accessible ‘Berkeley’ dataset provided with the *sitar* package.

The remaining subsections give an overview to using LME models with linear and natural cubic splines, and SITAR models to estimate population-average trajectories (and individual variation from these), and extensions to latent trajectory models to identify heterogenous trajectories in subpopulations. These four modelling approaches are summarised in **Table 1**.

**Table 1.** Overview of linear spline LME models, natural cubic spline LME models, SITAR, and latent trajectory models for analysing nonlinear growth trajectories of a single repeatedly measured continuous outcome.

|  | Linear spline LME model | Natural cubic spline LME model | SITAR | Latent trajectory model |
| --- | --- | --- | --- | --- |
| Description | linear mixed-effects model with a linear spline function of the independent time variable | linear mixed-effects model with a restricted cubic spline function of the independent time variable | nonlinear mixed-effects model based on the shape invariant growth model | heterogenous growth curves fit to unknown subgroups of individuals |
| Advantages | easy to interpret the spline slope coefficients; can describe growth rate during different periods of the growth process | continuous 1<sup>st</sup> & 2<sup>nd</sup> derivatives give smoother trajectory and can identify points of peaks/troughs; linearity constraint gives a more reliable trajectory shape as less erratic at the tails of distribution | has useful features of the natural cubic spline, easy to estimate individual growth features – most notably individual ages at peak growth velocity | can identify unobserved sub-groups of individuals sharing distinct growth trajectories if any exist |
| Limitations | biologically implausible sudden changes in velocity (i.e., at the knots); erratic at the tails; cannot identify points of velocity maxima/minima; position (and location) of knots important | coefficients difficult to interpret (so plotting is more useful); can be challenging to estimate the individual growth curves due to complex spline basis functions used by the statistical software | may not work well for complex growth patterns e.g., with multiple peaks and troughs or where the growth curve does not plateau in adulthood | difficult to identify the optimal number of sub-groups; may identify implausible subgroups; trajectories tend not to replicate in other cohorts |
| R package(s) | lme4; lspline | lme4; splines | sitar | lcmm, splines |

All models can include all individuals with at least one observed outcome measure, with valid estimates obtained under the assumption of outcome data missing at random (MAR) depending on observed values of the outcome and/or covariates. Note all models assume no autocorrelation (in our example, there are wide enough gaps between measures to assume that here).

### Linear spline LME models

A spline function is a set of piecewise polynomials that are joined together at points called knots [14]. Regression splines are a type of splines formed by a linear combination of the transformed time variable (e.g., age), thus they can be used within LME to model the repeated outcome as a nonlinear function of time. The simplest function is the linear spline where growth is described by a series of connected lines joined at knots, where the slope can change after each knot [3]. For example, a linear spline for age (measured from 5–40 years) with 2 knots at ages 10 and 15 years produces three different linear slopes of the repeated measure (e.g., weight): 5 to ≤10, 10 to ≤15, and 15 to ≤40 years.

The LME model in (1) can be rewritten to include a linear spline function for age *b*(*t*) with *K* knots *ξ*_1_ < *ξ*_2_ < ⃛ *ξ*_*K*_ as:

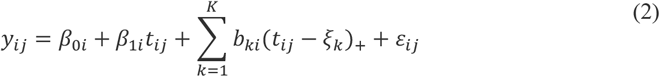

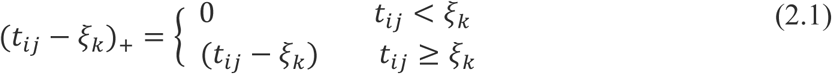

The model in (2) includes a linear spline in both the fixed and random effects (with *b*_*ki*_ having fixed effect *b*_*k*_ and random effect *v*_*ki*_) which gives nonlinear mean and individual trajectories, respectively. The fixed and random splines are assumed to have the same knots in (2) however, it is possible to allow fewer knots in the random spline. If the aim is solely to model a nonlinear mean trajectory, then (2) can be simplified by replacing the random spline with a random line – here, *β*_*oi*_ and *β*_1*i*_ have similar interpretation as in (1), with random effect *v*_*ki*_ omitted (i.e., *b*_*ki*_ = *b*_*k*_). Rate of change of a linear spline (1^st^ derivative) is not continuous at the knots. An alternative function with continuous 1^st^ and 2^nd^ derivatives is the natural cubic spline.

### Natural cubic spline LME models

A natural cubic spline (restricted cubic spline) is a set of cubic polynomials with continuity and slope constraints at each knot, and additional constraint of linearity at the extremes of the curve, before the first knot and after the last knot [11, 14-16]. This linearity constraint makes the trajectory less erratic at the ends of the distribution and so more reliable than linear splines (and unrestricted cubic splines), and more parsimonious for complex shapes than a linear spline with many knots.

An LME model that includes a natural cubic spline function *b*(*t*) with *K* knots *ξ*_1_ < *ξ*_2_ < ⃛ *ξ*_*K*_ (and a linearity constraint for values *t* < *ξ*_1_ and *t* > *ξ*_*K*_) can be written as:

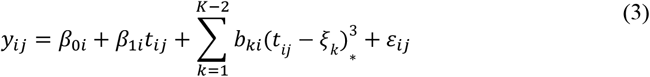

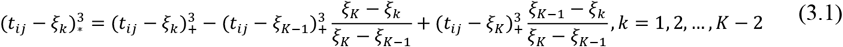

In words, a natural cubic spline for age (measured from 5–40 years) with 2 knots at 10 and 15 years (and with first and last knots at 5 and 40 years, respectively), invokes 3 cubic polynomials: between 5 to ≤10, 10 to ≤15, and 15 to ≤40 years, and has its curvature equal to 0 at ages 5 and 40 years. If the first and last knots were placed at older and younger ages respectively, then the curve would be linear from the first and last knot to the youngest and oldest ages, respectively. Note we have defined the natural cubic spline using truncated power basis (like how the linear spline was defined). The analysis software creates this function using a B-spline basis which is mathematically challenging to represent but numerically more stable. Whatever basis is used, if the polynomial degree and knots are identical, then the spline will always be the same.

### Choosing number and location of knots

The flexibility of regression splines is determined by the number/position of the knot points. A small number of knots (between 3 and 5 knots) provides a good fit to some patterns [15] though, with many repeats (e.g., data spanning many decades) more knots may be required. Approaches to selecting number/position of knots include (i) placing knots at quantiles of the age distribution using equally spaced knots, (iii) inspecting smoothing curves and using these to select knots, (iv) starting with many knots and reducing their number, and (v) placing knots at mean age of data collection [3]. For natural cubic splines, the number of knots (rather than their position) is more important [15]. Model selection can be done informally (inspecting plots from competing models). Valid comparison between models with different knots (non-nested mean structures) can be done using likelihood-based information criteria if maximum likelihood (ML) estimation is used [17]. Cross-validation is also useful for model selection [18]. If knot position was primary interest (e.g., testing sensitive periods), then topic knowledge can inform placement of knots [19].

### SITAR models

SITAR is a shape invariant nonlinear mixed-effects model [5, 20]. Whereas LME spline models are linear models that allow terms to describe a non-linear trajectory with age, nonlinear mixed-effects models are fundamentally nonlinear in the coefficients [11]. SITAR assumes that a study population has a common characteristic curve (fitted as fixed effects), which through shifting and scaling (by a set of 3 random effects) can be transformed into any individual curve. Following the notation in Cole *et al* [5, 21], a SITAR model for outcome *y* can be written as:

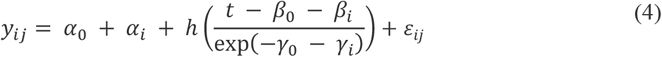

where *y*_*ij*_ is the outcome measurement for individual *i*at age *j*; *α*_0_, *β* _0_, *γ* _0_ are fixed effects; α_*i*_, *β*_*i*_, and *γ*_*i*_ are random effects for the *i*th individual; *h*(*t*) is a natural cubic spline curve; and *ε*_*ij*_ are independent normally distributed errors. The 3 random effects describe the size (*α*_*i*_), timing (*β*_*i*_), and intensity (*γ*_*i*_) of individual growth relative to the mean growth curve. *α*_*i*_ adjusts for the differences in *y* and geometrically reflects individual shifts up or down (translation) in the mean curve; *β*_*i*_ adjusts for differences in the timing of peak growth in *y* and geometrically reflects left to right shifts (translation) in the mean curve; and *γ*_*i*_ adjusts for the duration of the growth spurt and geometrically corresponds to shrinking or stretching of the age scale and rotating the curve.

Note, a key difference from other mixed effects models, with or without a natural cubic spline mean curve, is that SITAR models growth on both the x-and y-axes – this allows differences in developmental age to be modelled. Common practice in selecting the best fitting SITAR models is to compare models with varying number of knots placed at quantiles of the age distribution for the spline curve [5]. The internal SITAR model structure is also customisable [22].

### Latent trajectory models

The spline and SITAR models described above assume that the population is homogenous and described at the population level by a mean trajectory, with variability of individuals about this mean. As an alternative, latent trajectory modelling assumes there is a heterogenous population composed of unknown subgroups (latent classes) of individuals, each characterised by a unique mean trajectory profile [6, 23, 24]. These models aim to minimise within group variance and maximise between group differences so that individuals are more similar within groups than between groups. Each individual has a probability of belonging to each latent class and is assigned to the class with the highest probability. Class membership is defined using a latent discrete random variable, and membership probability described by a multinomial logistic model.

Several modelling approaches are possible (see [6] for a recent overview). These include models that ignore the longitudinal structure (referred to as longitudinal latent class analysis) and models with no variability between individuals within subgroups (known as latent class growth analysis or group-based trajectory models). A direct extension of standard mixed-effects models is growth mixture models, which involves fitting multiple growth curves to subgroups of individuals that share a common trajectory. Following the notation in Herle *et al* [6], the LME model in (1) can be rewritten as a growth mixture model as:

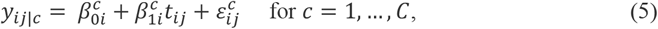

where *C* indicates number of latent classes, with probabilities *p*_*c*_,*c* = 1, …, *C*, with 0 ≤ *p*_*c*_ ≤ 1 and 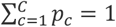. All other terms are defined as before but specifically for each class. Growth can be parameterised as nonlinear, e.g., using a natural cubic spline curve in each class (assuming same number/position of knots). Class-specific covariances for individual-level error terms can be included, and fixed and random effects can be class specific.

Model estimation is conditional on a pre-specified number of classes, with the optimal number of classes identified through a combination of approaches. These include assessing interpretability and plausibility of classes e.g., inspecting if trajectories show biologically plausible patterns and examining characteristics (e.g., socioeconomic position) of the classes [25], information criteria, entropy (statistic for class separation), and numerically meaningful sub-groups (e.g., ≥5% class size). Models with >1 class are prone to local maxima solutions (convergence to best solution in a neighbourhood of the parameter space, rather than the global maximum (largest loglikelihood)). This can be avoided by using different starting values [26].

## ILLUSTRATIVE EXAMPLE

In this section we use data from three cohort studies to demonstrate how the four approaches can help answer our research aim to characterise bone mineral content (BMC) growth trajectories and their sex differences.

### Bone mass through the life-course

Bone mass in early life is thought to be an important determinant of fracture and osteoporosis risk in later life [27] however, few studies have described its developmental trajectory. Furthermore, sex differences in osteoporotic fracture are assumed to be due to menopause but may also reflect early sexual dimorphism in bone development. This is a timely exploration, given the availability of studies with repeated measurements of BMC (a marker of bone strength).

### Studies and measurements

Three studies with repeated BMC measurements from childhood to adulthood were included (**Table 2, Online Resource 1**): Avon Longitudinal Study of Parents and Children (ALSPAC) [28, 29], Bone Mineral Density in Childhood Study (BMDCS) [30], and Pediatric Bone Mineral Accrual Study (PBMAS) [31]. Total-body (excluding-head) BMC was measured in grams using whole-body Dual-Energy X-ray Absorptiometry (DXA) scans. Of note, the studies used DXA devices from different manufacturers (Lunar vs. Hologic) which scale differently and are not interchangeable but repeat scans within studies were acquired on the same device. Individuals from each study were included if they had ≥1 measure of BMC and no missing data on age at DXA scan (in years) or sex. Analyses were restricted to white ethnicity because 2 cohorts were ethnically homogeneous [29, 31]. The final analysis samples comprised 3,888 males and 4,007 females in ALSPAC, 465 males and 488 females in BMDCS, and 112 males and 127 females in PBMAS. All studies had ethics approval and obtained parental or participant informed consent.

**Table 2.** Characteristics of the three cohort studies included in the trajectory modelling

|  |  |  |  |
| --- | --- | --- | --- |
| Study name | Avon Longitudinal Study of Parents and Children (ALSPAC) | Bone Mineral Density in Childhood Study (BMDCS) | Pediatric Bone Mineral Accrual Study (PBMAS) |
| Design | birth cohort study (started in 1990-1992) | child cohort study (started in 2002-2003) | child cohort study (started in 1991) |
| Region and country | catchment area of 3 health authorities in Southwest England, UK | 5 USA clinic centres: Los Angeles, New York, Cincinnati, Omaha, Philadelphia | 2 elementary schools, Saskatoon, Saskatchewan, Canada |
| Birth years | 1990-1992 | 1985-1997 | 1983-1976 |
| Ethnicity | 98% white ethnicity | ethnically diverse | 95% white ethnicity |
| DXA device used to measure BMC | Lunar Prodigy | Hologic QDR-4500A | Hologic QDR-2000 |
| Mean age at the baseline/youngest DXA scan (range) | 9.9 years (8.8-11.7 years) | 10.8 years (6.0-17.0 years) | 11.8 years (8.0-15.1 years) |
| Mean age at last/oldest DXA scan (range) | 24.6 years (22.4-26.5 years) | 16.1 years (6.9-23.3 years) | 37.3 years (34.3-40.2 years) |
| Frequency and the maximum number of repeated DXA scans | up to 6 repeated scans at mean ages 9.9, 11.7, 13.8, 15.4, 17.8, and 24.6 years | up to 7 yearly repeated scans | Up to 16 repeated scans (1991-1998, 2003-2005, 2007-2011 and 2016-2017) |
| Individuals included in the analysis* | 4007 females<br>3888 males | 488 females<br>465 males | 127 females<br>112 males |

\*Trajectory modelling was restricted to white ethnicity individuals.

### Statistical analysis

Analyses were performed in R version 4.0.2 (R Project for Statistical Computing) and RStudio version 1.3.1 (RStudio Team). R code is available at https://github.com/aelhak/nltmr/. Synthetic versions of the PBMAS cohorts were simulated from natural cubic spline LME models [32] and can be found in the same repository.

Prior to trajectory modelling scatterplots of BMC against age, and line plots of the individual BMC trajectories were used to inspect the form of the trajectory and identify clearly outlying observations. The datasets used for trajectory modelling **(Fig 2)** showed as expected nonlinear change in BMC with age, and higher BMC in ALSPAC due to Lunar device. The numbers of individuals at each visit were described and age and BMC were summarised with means and standard deviations (**Online Resource 1**). Models were fitted separately by sex due to expected difference in bone accrual and our aim was to explore this in the illustrative example.

**Fig. 2.**
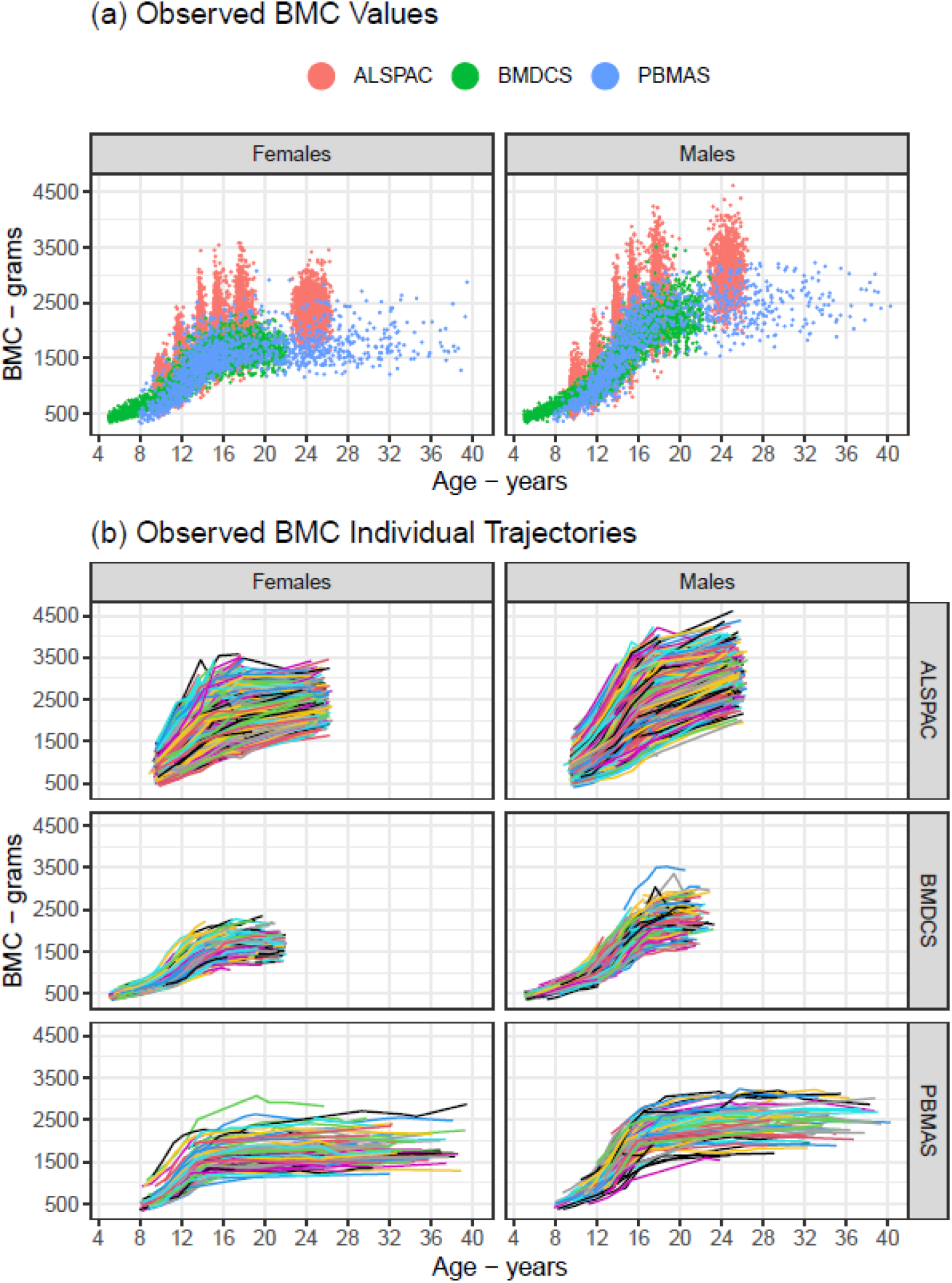
Plots of the cohort datasets used in the trajectory modelling showing (a) bone mineral content (BMC) values at each age, and (b) BMC individual trajectories.

Linear and natural cubic spline LME models were fitted using the ‘*lme4’* package [33]. Models with 2 to 6 knots (at quantiles of the age distribution) in the fixed effects curve were compared. Nonlinear individual trajectories were allowed by including random effects spline with 1 knot at the median. SITAR models with 2 to 6 knots (at quantiles of the age distribution) in the spline curve (and three random-effects) were fitted using the ‘*sitar’* package [22]. LME and SITAR models were fitted by ML estimation and best fitting models (optimum number of knots) were determined by smallest Bayesian Information Criterion value (BIC0 (**Online Resource 2**).

Goodness of fit for selected models was assessed by examining residuals (conditional on the random effects) from LME models and variance in BMC explained by SITAR models. The selected models were used to describe BMC growth trajectory and growth velocity. Slopes of the fixed effects spline segments from linear spline models were used to summarise mean BMC velocity during different age windows and identify windows for peak growth. Mean peak and age at peak velocity were obtained from natural cubic spline LME and SITAR models by differentiating mean spline curves.

Growth mixture models were fitted using the ‘*lcmm’* package [26]. The forms of the best-fitting mean natural cubic spline curves were used to model the fixed effects age curve. Models included random intercepts and random linear age slopes. Models with different numbers of latent classes were compared: from a 1-class model (i.e., a standard LME model where all individuals follow a single mean trajectory) up to models with 5 classes. Models with >1 class included class-specific random effects covariance matrices. An automatic search procedure was used to estimate each 2-5 class model for 100 iterations using random initial values from the distribution of the 1-class model. Optimal number of classes was chosen by inspecting predicted trajectory sub-groups from each model for biological plausibility, in addition to the smallest BIC and biggest entropy, and by excluding small class size (≥5%). Goodness of fit and discrimination capacity of the selected models was assessed by calculating posterior class membership probabilities [26].

## Results

The mean predicted BMC trajectories in each cohort from the linear spline LME (**Fig. 3**), natural cubic spline LME (**Fig. 4**), and SITAR models (**Fig. 5**) showed that BMC increased with age up to a plateau in young adulthood, and thereafter remained stable to age 40 (PBMAS). All models showed evidence of steeper growth trajectories through adolescence coinciding with emerging sex differences, with males subsequently having higher BMC than females, and plateauing later than females. BMC trajectories from all models were broadly similar (**Fig. 6**). Both spline LME models and SITAR provided a good fit to the data (**Online Resource 2**). Mean BMC growth velocity in different age windows (from linear spline LME models) peaked during adolescence, and the peak was lower and occurred earlier in females than males (**Table 3**). Mean BMC growth velocity curves from natural cubic spline LME and SITAR models were similar (**Fig. 4-5**): for all cohorts, mean ages at peak velocity from both models were within the age windows for peak growth identified by the linear spline models (**Table 4**).

**Fig. 3.**
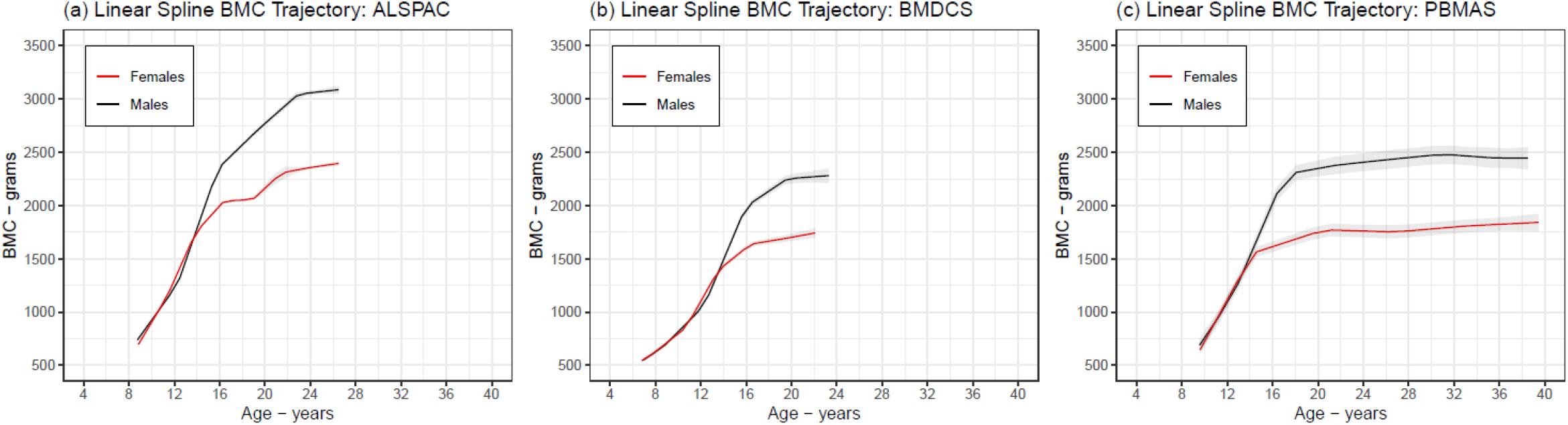
Mean BMC growth trajectory from the selected linear spline LME models. Shaded areas around the mean trajectories represent 95% confidence intervals.

**Fig. 4.**
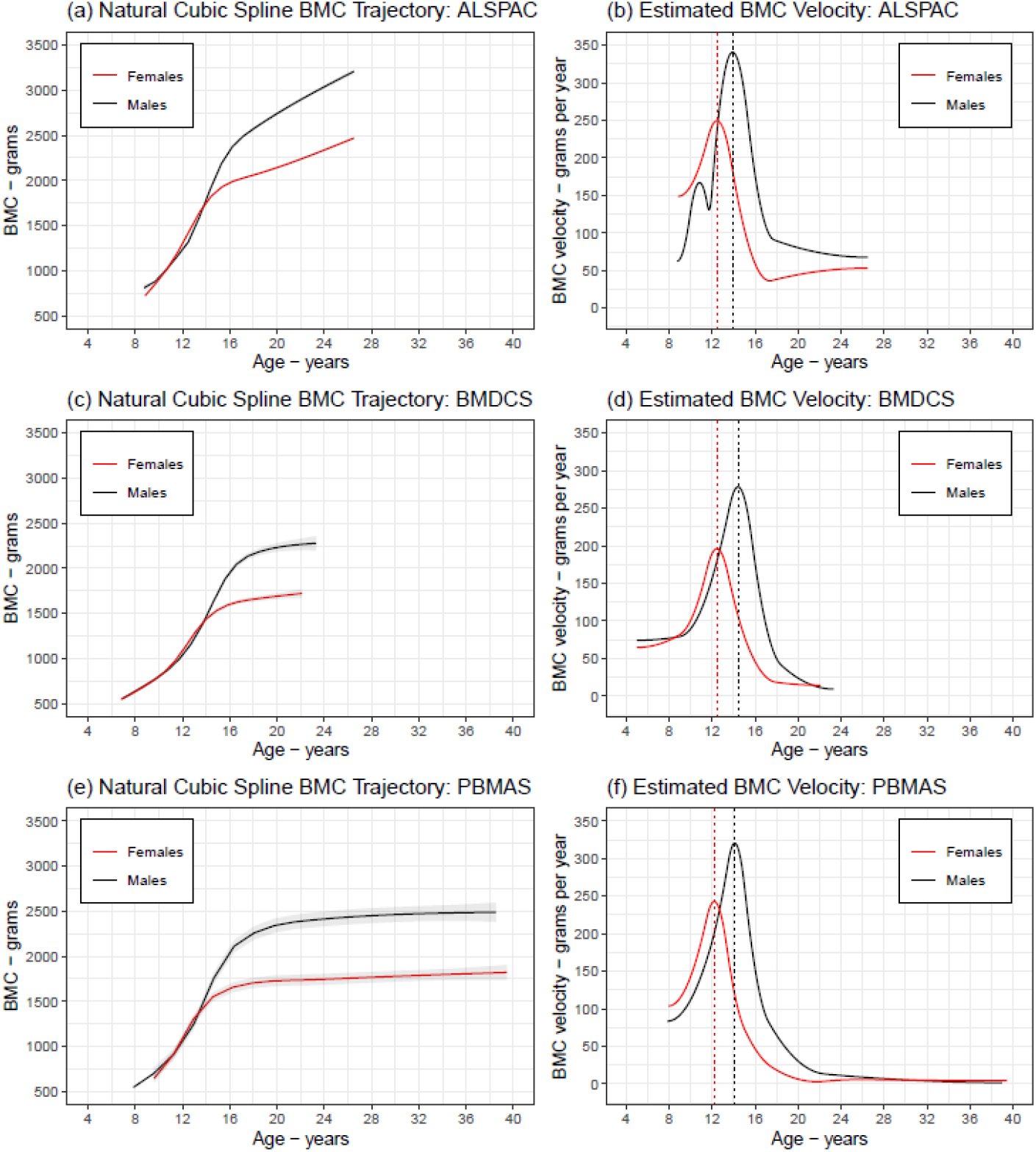
Mean BMC growth trajectory (left panels) and mean BMC growth velocity and age at peak velocity (right panels) from the selected natural cubic spline LME models. Shaded areas around the mean growth trajectories represent 95% confidence

**Fig. 5.**
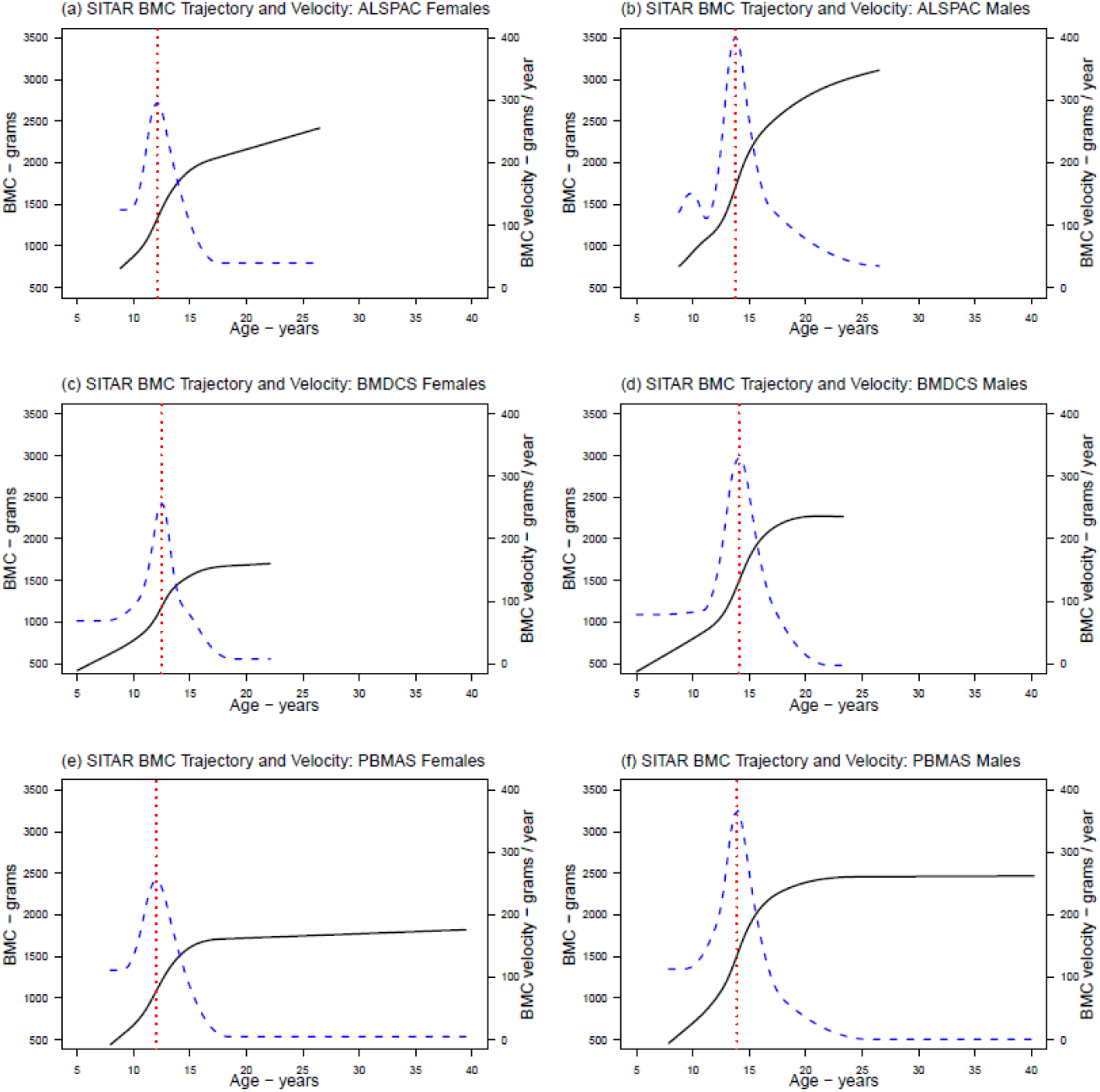
Mean BMC growth trajectory (solid black curves), mean BMC growth velocity (dashed blue curves), and mean age at peak BMC velocity (vertical red lines) from the selected SITAR models.

**Fig. 6.**
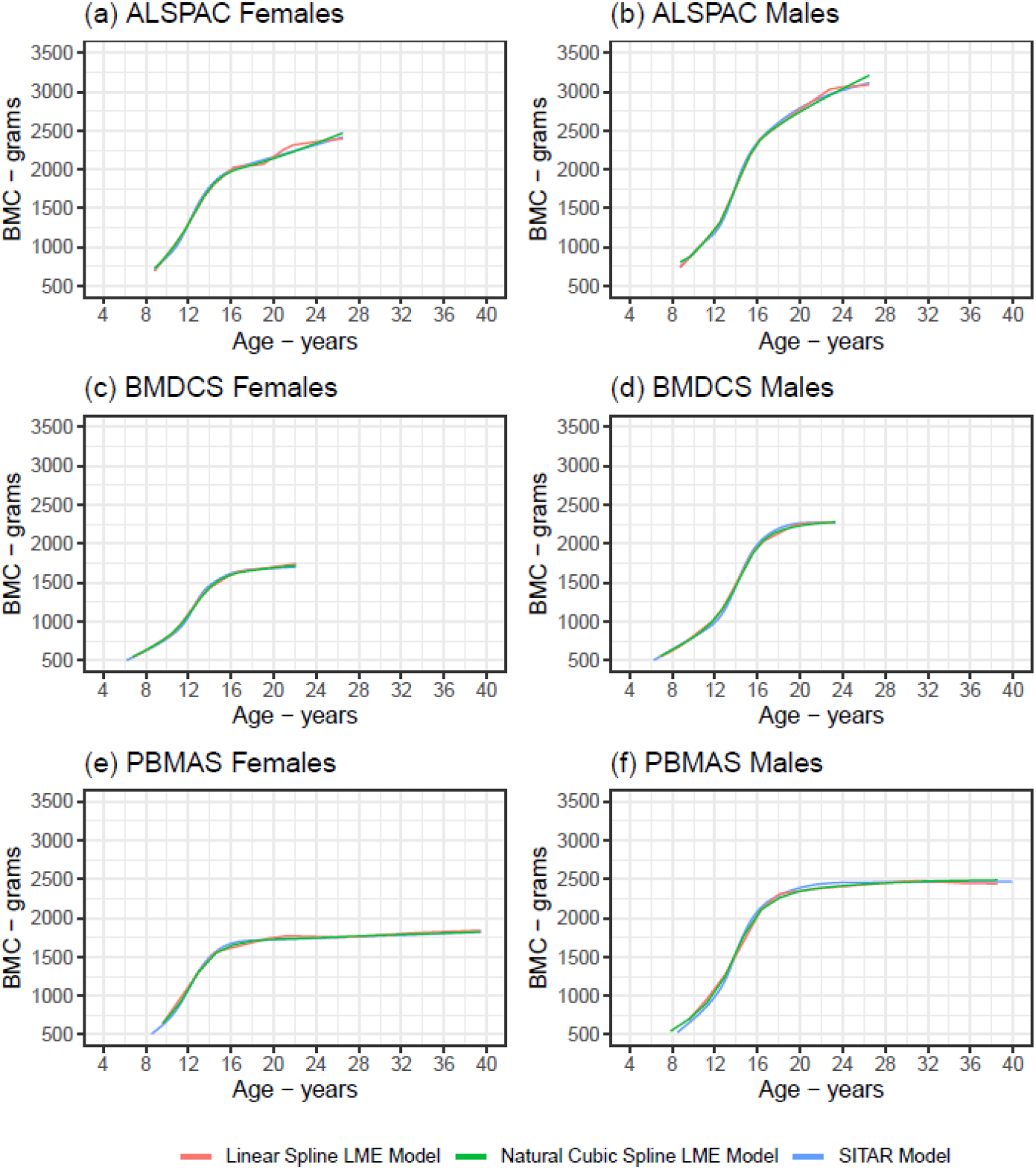
Overlayed mean BMC growth trajectories from the selected linear spline LME models, natural cubic spline LME models, and SITAR models.

**Table 3.** Estimated mean BMC growth velocity during different age windows from the selected linear spline LME models.

|  | Grams per year change in BMC [(mean (95% CI)] |  |  |
| --- | --- | --- | --- |
|  | ALSPAC | BMDCS | PBMAS |
| *Females* |  |  |  |
| 8.8y to 11.4y (ALSPAC); 5.0y to 7.9y (BMDCS); 7.9y to 14.2y (PBMAS) | 177.7 (173.8 to 181.6) | 67.6 (60.1 to 75.1) | 195.4 (189.7 to 201.1) |
| 11.4y to 13.9y (ALSPAC); 7.9y to 10.7y (BMDCS); 14.2y to 20.5y (PBMAS) | 238.9 (236.0 to 241.7) | 84.1 (79.0 to 89.2) | 34.7 (30.0 to 39.5) |
| 13.9y to 16.4y (ALSPAC); 10.7y to 13.6y (BMDCS); 20.5y to 26.9y (PBMAS) | 117.3 (113.2 to 121.3) | 189.0 (184.7 to 193.3) | -3.3 (-7.5 to 0.9) |
| 16.4y to 18.9y (ALSPAC); 13.6y to 16.4y (BMDCS); 26.9y to 33.2y (PBMAS) | 5.0 (-1.3 to 11.3) | 85.7 (80.1 to 91.3) | 9.1 (3.0 to 15.1) |
| 18.9y to 21.5y (ALSPAC); 16.4y to 19.3y (BMDCS); 33.2y to 39.5y (PBMAS) | 99.2 (74.2 to 124.2) | 17.4 (10.6 to 24.1) | 5.2 (-5.9 to 16.3) |
| 21.5y to 24.0y (ALSPAC); 19.3y to 22.1y (BMDCS) | 20.1 (-7.0 to 47.2) | 19.5 (11.1 to 27.8) | - |
| 24.0y to 26.5y (ALSPAC) | 15.2 (3.0 to 27.5) | - | - |
| *Males* |  |  |  |
| 8.8y to 12.3y (ALSPAC); 5.0y to 8.6y (BMDCS); 7.8y to 12.5y (PBMAS) | 148.5 (144.2 to 150.8) | 70.2 (61.7 to 78.7) | 157.5 (144.2 to 170.8) |
| 12.3y to 15.9y (ALSPAC); 8.6y to 12.3y (BMDCS); 12.5y to 17.1y (PBMAS) | 305.6 (302.5 to 308.8) | 107.4 (101.7 to 113.1) | 247.1 (238.7 to 255.4) |
| 15.9y to 19.4y (ALSPAC); 12.3y to 16.0y (BMDCS); 17.1y to 21.7y (PBMAS) | 103.3 (98.6 to 108.0) | 253.2 (248.4 to 258.0) | 19.3 (8.9 to 29.7) |
| 19.4y to 23.0y (ALSPAC); 16.0y to 19.6y (BMDCS); 21.7y to 26.3y (PBMAS) | 93.5 (84.1 to 102.8) | 71.4 (63.3 to 79.5) | 11.2 (0.8 to 21.6) |
| 23.0y to 26.5y (ALSPAC); 19.6y to 23.3y (BMDCS); 26.3y to 31.0y (PBMAS) | 11.7 (-6.4 to 29.9) | 8.0 (-3.5 to 19.6) | 10.7 (-1.5 to 22.8) |
| 31.0y to 35.6y (PBMAS) | - | - | -7.9 (-26.4 to 10.7) |
| 35.6y to 40.2y (PBMAS) | - | - | -0.4 (-32.7 to 32.0) |

Age windows are defined by the number and position of the knots, which were placed at quantiles of the age distribution.

**Table 4.** Estimated mean age at peak BMC velocity from the selected natural cubic spline LME models, and selected SITAR models. For comparison, the age windows for peak BMC velocity from the linear spline LME models are also presented.

|  | Females |  |  |  | Males |  |  |
| --- | --- | --- | --- | --- | --- | --- | --- |
|  | ALSPAC | BMDCS | PBMAS |  | ALSPAC | BMDCS | PBMAS |
| *Mean age at peak BMC velocity (years)* |  |  |  |  |  |  |  |
| Natural cubic spline LME model | 12.5 | 12.4 | 12.2 |  | 13.9 | 14.4 | 14.1 |
| SITAR model | 12.1 | 12.5 | 12.0 |  | 13.8 | 14.1 | 13.9 |
| *age window for peak BMC velocity from the linear spline LME model (years)* | 11.4 to 13.9 | 10.7 to 13.6 | 7.9 to 14.2 |  | 12.3 to 15.9 | 12.3 to 16.0 | 12.5 to 17.1 |

The selected growth mixture models identified 3 subgroups for females and 2 subgroups for males in ALSPAC and BMDCS, and 4 for females and 3 for males in PBMAS (**Fig. 7)**. Overall, differences in mean BMC between subgroups were larger during adolescence than in childhood and adulthood. One group with 15% of PBMAS females had higher mean BMC up to age 40 than the remaining three groups. A group comprising 27% of PBMAS males reached a lower peak and showed signs of bone loss by age 40, compared to the other two groups. Model discrimination capacity was better in PBMAS (and BMDCS) than ALSPAC, likewise entropy was high in PBMAS and low in ALSPAC and BMDCS (**Online Resource 3**).

**Fig. 7.**
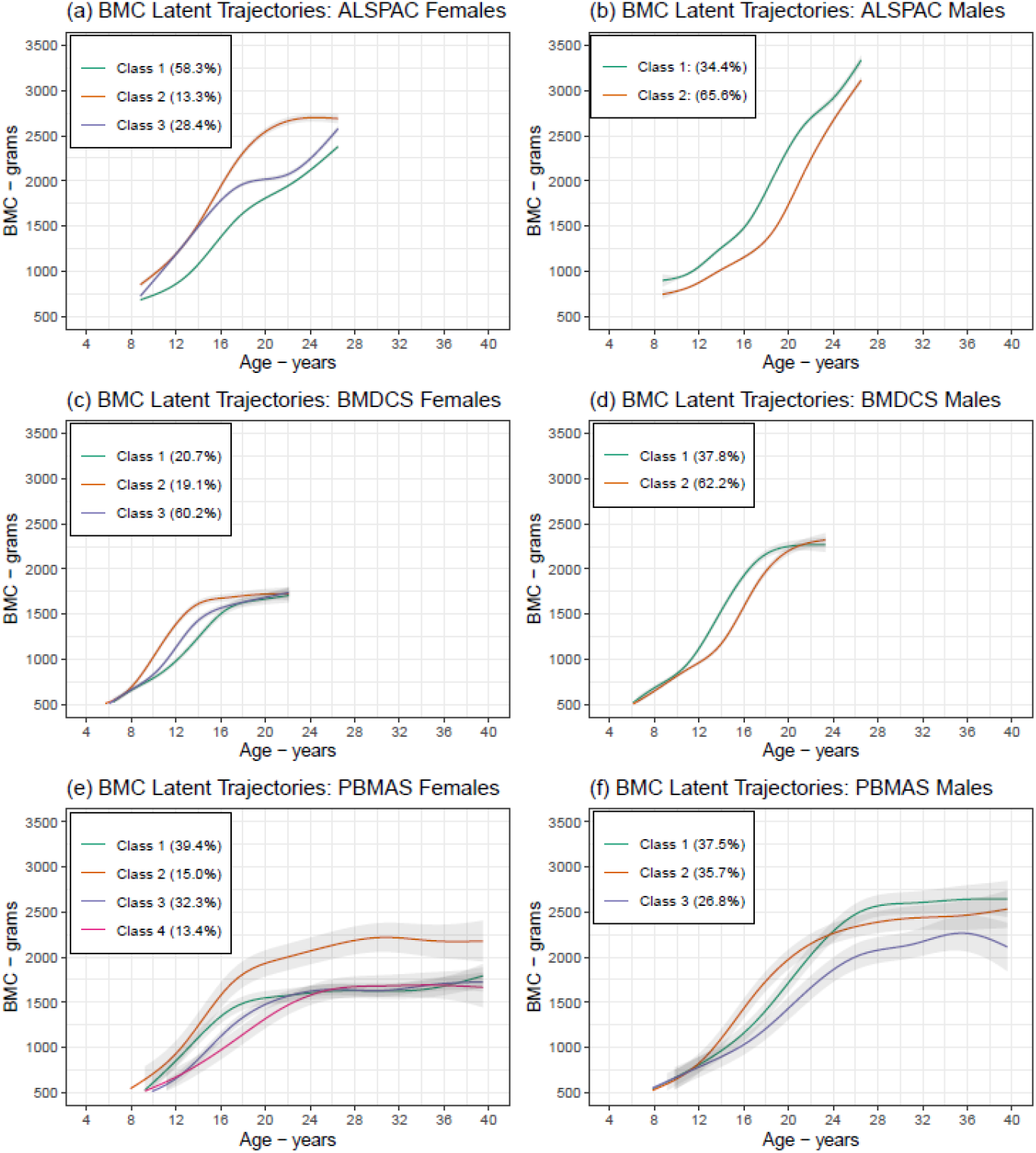
Mean BMC growth trajectories by subgroup (latent class) from the selected growth mixture models. Colours distinguish between latent trajectory subgroups within subplots and should not be used to compare between subplots. Shaded areas around the mean trajectories represent 95% confidence intervals. The numbers in each class are: ALSPAC females (class 1: n=2337, class 2: n=531, class 3: n=1139), ALSPAC males (class 1: n=1339, class 2: n=2549), BMDCS females (class 1: n=101, class 2: n=93, class 3: n=294), BMDCS males (class 1: n=176, class 2: n=289), PBMAS females (class 1: n=50, class 2: n=19, class 3: n=41, class 4: n=17), PBMAS males (class 1: n=42, class 2: n=40, class 3: n=30).

### Interpretation

Our results provide evidence on bone mineral accrual from 5-40 years. LME splines and SITAR models showed that the levels and rates of change in BMC were greater for males than females, with peak gains in adolescence in both, but later in males than females. Growth mixture models identified potentially distinct trajectory sub-groups, with the greatest between-group differences seen in adolescence. Whilst the main aim of these analyses is to illustrate different modelling approaches, our results are consistent with previous studies on sex differences in BMC and suggest that in both sexes puberty is an important period for peak bone accrual [34-36].

## DISCUSSION

### Choosing a modelling approach

All the modelling techniques presented may be used to model BMC and other growth processes. Choice of method will be determined by research question, including whether this is concerned with differences in mean change over time (e.g., with LME and SITAR models) or whether the aim is to identify data-driven subgroups for patterns of change), complexity of the underlying trajectory, and data availability i.e., number of individuals and repeat measurements (see [37] for a discussion on sample size in growth models). There may also be value in using a combination of approaches [38], like in this paper. Both spline LME models and SITAR were previously used on height/weight/BMI [4, 21, 39, 40]; linear splines and SITAR were previously used to model bone density [34, 41]; linear and natural cubic spline LME were used to analyse blood pressure change [42, 43]. All models in this paper can handle unbalanced datasets (i.e., with individually varying measurement occasions, as in our illustrative example).

When the main aim is to quantify growth rate at different periods of the life-course, then linear splines may be preferred because of their more interpretable slope coefficients compared to the natural cubic spline LME and SITAR models. If the aim is to describe the shape of the trajectory or identify specific peaks and troughs (e.g., age at peak velocity), then natural cubic spline LME or SITAR are more useful, because linear spline cannot identify points of maxima/minima (but can identify periods/age windows). The number and spacing of repeated measures can influence model convergence and the complexity that can be allowed [44, 45]. SITAR was designed to model adolescent growth in height, and (like other nonlinear models) its parameters reflect its specific purpose. Hence, SITAR may not work well for complex trajectories (e.g., depressive symptoms [46]), and the natural cubic splines LME may offer more flexible alternatives. Note however, SITAR fitted without the timing fixed effect is analogous to a random intercept random slope model, and so should be at least as flexible as LME. Other strategies that may improve the SITAR model include logging age or outcome and manually specifying the knots.[44, 47].

Latent trajectory models have been used to identify trajectory subgroups for BMI [48], depressive symptoms [49], physical activity [50], glucose response [51], and environmental exposures [52], among others. Growth mixture models and latent trajectory models in general take longer to run, particularly in larger cohorts. Less complex group-based trajectory models are faster; however, subgroup differences may just reflect within class variability, which is likely to be absorbed by the random effects in growth mixture models. If exploring specific hypotheses (e.g., sensitive periods), clustering (data driven) approaches might not provide a suitable sub-group to test this, e.g., if the aim was to explore the hypothesis that lower birth weight followed by faster growth in the first 1000 days increased cardiovascular risk, this approach might not identify a latent class with this specific growth pattern. It is difficult to determine the optimal number of sub-groups, including whether the latent classes are meaningful or if the model is splitting the distribution of the random effects into a larger group and smaller extremes. Model selection is often subjective, and trajectory subgroups tend to be cohort-specific and do not replicate in other cohorts – so it is essential to follow reporting guidelines [24].

### Identifying determinants and outcomes of trajectories

The models in this paper can be extended to include early life exposures and later outcomes to explore their associations with trajectories. Choice of method will depend on the research aims. Exposure variables can be added within LME spline models as fixed effects and as interactions with splines to test their effect on trajectories and growth rate [3, 34, 53]. Individual growth features (e.g., peak velocity and age at peak velocity) can be obtained from spline LME model random effects and used in separate analyses as outcomes or exposures [4, 54] – however, it is important to have enough complexity in the random-effects splines for sufficient between-person variability (**Fig 8**). Individual growth features are easily obtained from SITAR models and can be used in subsequent analyses to examine associations with exposures or outcomes [41, 55]. Of note, 2-stage approaches may be more biased than 1-stage joint modelling [56, 57]. Exposures and outcomes can be related to latent trajectories in a joint model or a multistage process where the subgroups are first identified and then used in separate models (unweighted or (preferably) weighed for classification probabilities) to examine associations [26, 48-52]. If the aim was to identify effects of repeated exposure, then a structured modelling approach may be useful for testing competing hypotheses [58].

**Fig. 8.**
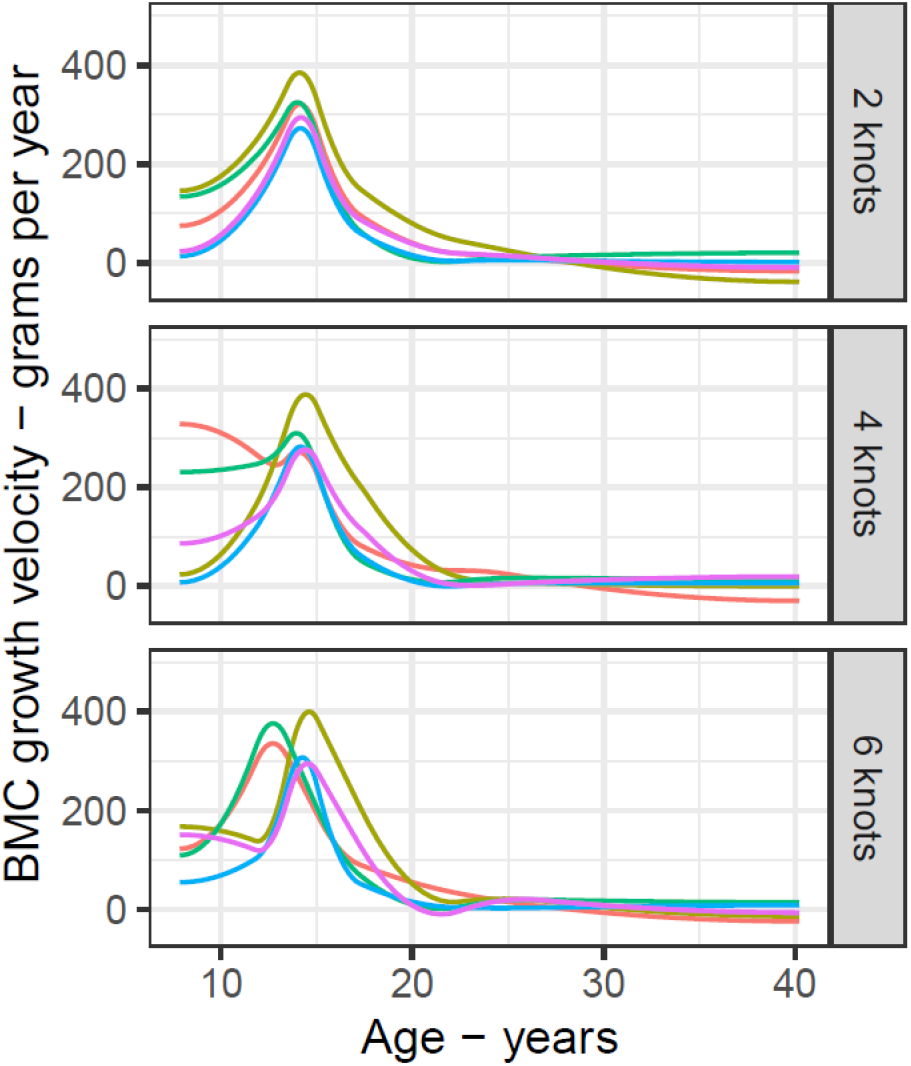
Effects of increasing the number of knots in the random effects spline on the individual growth velocity curves from the natural cubic spline LME model. Plot shows the velocity curves for 5 randomly selected individuals, obtained from natural cubic spline LME models in PBMAS males with 2, 4, and 6 knots for the random effects spline curve. All models included 6 knots in the fixed effects spline curve. All knots were placed at quantiles of age distribution.

It is important to identify potential biases and explore ways for mitigating them when analysing (causal) associations between trajectories and exposures or outcomes in cohort studies. Missing data can bias associations depending on mechanism, and appropriate approaches to describe and handle missing data should be explored [11, 59-63]. In a repeated measure setting, individuals with missing outcome values can be included in the estimation sample if they have at least one observed outcome value. Mixed-effects models give unbiased results (i.e., not biased by missing data) when the probability that an outcome value is missing depends on observed values of the outcome (i.e., outcome missing at random (MAR) depending on observed outcome values). Bias due to missing data will occur for these models when the probability that the outcome is missing depends on underlying missing values (i.e., outcome missing not at random (MNAR) depending on missing outcome values). With incomplete covariates, bias can occur when the probability of excluding an individual with missing covariate data is related to the outcome. Regardless of bias, excluding individuals with missing covariate information will often mean discarding useful observed data, leading to imprecision [64].

Confounding can lead to spurious associations between trajectories and exposures or outcomes. Confounders (factors causally related to exposure and outcome) should be identified (e.g., using Directed Acyclic Graphs) and controlled for, taking care not to adjust for mediators (factors on the causal pathways) [65, 66]. Even with adjustment, residual confounding (from using poorly measured confounders or not adjusting for important confounders) can bias results. Useful strategies for checking if residual confounding influences results include negative control variables and comparing cohorts with different confounding structures [38, 67-69].

### Comparing and modelling trajectories across cohorts

It is important to be aware of potential differences in the participants, data collection methods and analysis models when comparing cohort-specific trajectories from different studies. For example, the higher BMC due to the Lunar machine in ALSPAC means it is inaccurate to conclude that Britons had higher BMC (and peak velocity) than North Americans. Another example is effect of medication use by older cohorts on blood pressure trajectories [70]. Multicohort collaborations provide unique opportunities to jointly model trajectories across different cohorts and extend amounts of the life-course studied [71]. However, this creates additional challenges including on model selection and missing data (see [72] for a recent discussion of challenges and solutions to multicohort modelling). Whatever approach is taken (cohort-specific or multicohort modelling), data harmonisation is an important first step that involves making data comparable across studies [73], e.g. DXA reference standards to harmonise BMC [74]. Because age was fully harmonised in our example, a simple approach to obtaining valid pooled estimates of age at peak velocity is fitting a growth model to all individuals with BMC expressed in cohort-specific SD units (**Fig 9**).

**Fig. 9.**
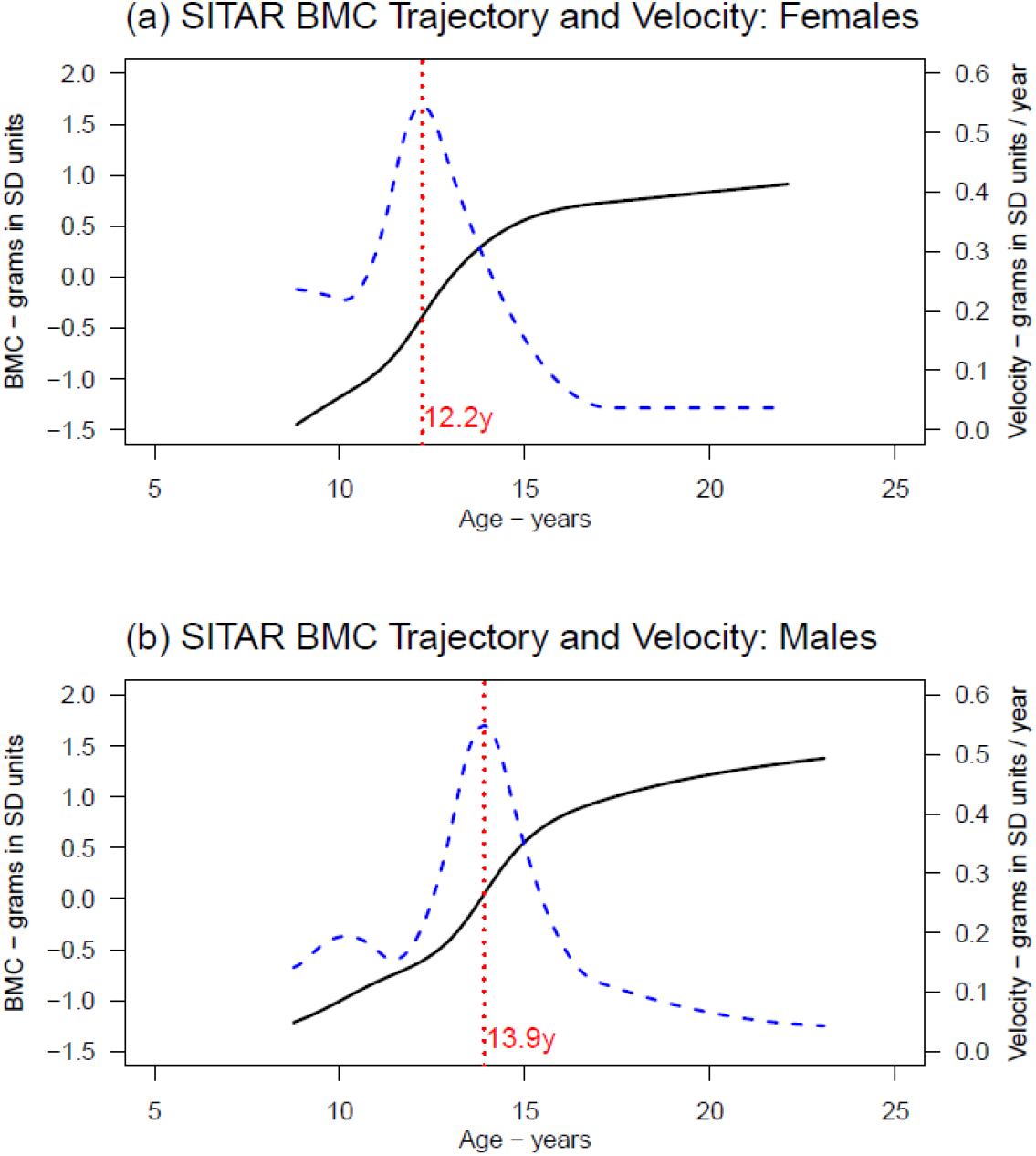
Pooled mean BMC growth trajectory (solid black curves), mean BMC growth velocity (dashed blue curves), and mean age at peak BMC velocity (vertical red lines) from SITAR models applied to individual participant data (ALSPAC, BMDCS and PBMAS). Sex-specific individual participant data SITAR models were fitted to ALSPAC, BMDCS and PBMAS combined, to obtained pooled estimates of the timing of peak BMC growth. This analysis included individuals with overlapping measurements (8.8 to 22.1 years) from the 3 cohorts (n=4431 for females and n=4359 for males.) To mitigate the cohort differences in BMC (higher values in ALSPAC due to Lunar machine), we modelled BMC in cohort-specific standardised units (mean=0 and SD=1), and the models were adjusted for cohort (as a fixed effect). Note it is not advised to fit SITAR to SD units as this distorts the underlying biology – though in our example, results are consistent with cohort-specific natural unit results.

### Conclusion

LME models with linear and natural cubic splines, SITAR, and growth mixture models are useful for describing nonlinear growth trajectories in longitudinal population studies. Choice of method depends on research aims, complexity of the trajectory, and available data. This illustrative paper and accompanying analysis code and example datasets will we hope be a useful resource for researchers interested in modelling nonlinear longitudinal trajectories.

## Data Availability

Details of all available data and the processes and procedures involved in accessing the ALSPAC resource can be found in the ALSPAC study website, which includes a fully searchable data dictionary and variable search tool (http://www.bristol.ac.uk/alspac/researchers/our-data/). The BMDCS data can be accessed through the NICHD DASH website (https://dash.nichd.nih.gov/). Researchers interested in accessing the PBMAS data should contact Professor Adam DG Baxter-Jones.

https://dash.nichd.nih.gov/

## Code availability

All the analysis code used in this paper can be found at https://github.com/aelhak/nltmr/.

## Authors’ contributions

AE developed the idea for the paper with initial input from DAL and with further development from all authors. AE conducted the analysis and wrote the first draft of the manuscript with initial input from DAL. All authors contributed to the development of the draft to its final version.

## Ethics approval

The ALSPAC study obtained ethics approval from the ALSPAC law and ethics committee and the local National Health Service research ethics committee. The BMDCS study was approved by the Institutional Review Board of each respective clinical center. The PBMAS study was approved by the University of Saskatchewan’s biomedical review committee.

## Acknowledgments

We thank the study participants from each cohort who took part in this study and the scientific and data collection teams from the ALSPAC, BMDCS and PBMAS cohorts. We are extremely grateful to all the families who took part in the ALSPAC study, the midwives for their help in recruiting them, and the whole ALSPAC team, which includes interviewers, computer and laboratory technicians, clerical workers, research scientists, volunteers, managers, receptionists and nurses. We acknowledge the BMDCS Principal Investigators: Vicente Gilsanz, MD, PhD, Heidi Kalkwarf, PhD, Joan Lappe, PhD, Sharon Oberfield, MD, John Shepherd, PhD, and the National Institute of Child Health and Human Development (NICHD) Data and Specimen Hub (DASH) for providing the BMDCS data that was used for this research.

## SUPPLEMENTARY MATERIAL

**Online Resource 1** Cohort characteristics and participant numbers, and age and BMC at each visit.

**Online Resource 2** BIC and fit statistics for linear spline LME models, natural cubic spline LME models, and SITAR models with 2 to 6 knots in the fixed effects spline curve

**Online Resource 3** Fit statistics and predicted mean BMC latent trajectories for growth mixture models with 1 to 5 latent classes.

